# Integrative Mendelian Randomization approaches for therapeutic target prioritisation in immune-mediated diseases

**DOI:** 10.1101/2024.04.27.24306475

**Authors:** Maria K. Sobczyk, Tom R Gaunt

## Abstract

**Background:** Immune-mediated diseases (IMD) encompass a wide range of autoimmune and inflammatory disorders with aetiology related to immune system dysfunction, signifying a disease area with great potential for drug repurposing. In this study, we employed the genetically informed Mendelian Randomization (MR) method with two distinct exposure types: immune blood cell abundance and protein quantitative trait loci (pQTL) to validate and repurpose 834 drug targets which have been investigated for IMD treatment.

**Methods:** Utilizing two-sample MR, we first established causal relationships between major peripheral immune cell types and 14 IMD. Robust associations, particularly with eosinophils, were confirmed across diseases such as asthma, eczema, sinusitis, and rheumatoid arthritis, revealing 59 high-confidence relationships. Intragenic variants associated with causal immune cell types were then extracted to create instruments for 371 existing IMD drug targets ("intermediate trait" MR). In parallel, we leveraged four large blood plasma protein QTL datasets to obtain complementary instruments for 361 targets ("pQTL" MR).

**Results:** In the intermediate trait MR analysis, we identified 811 gene-IMD associations (p-value <0.05), 169 of which were supported by strong colocalisation evidence (PP_H4_ ≥ 0.8). In the pQTL MR analysis, we similarly found 841 protein-IMD associations (p-value <0.05), 83 of which were confirmed with colocalization. Comparison with a list of approved drugs indicated low sensitivities across disease outcomes for both exposure types (intermediate trait MR: 0.49 ± 0.23 SD, pQTL MR: 0.28 ± 0.12 SD).

**Conclusions:** Drug targets identified in the pQTL and intermediate trait MR analyses show limited overlap (13%), presenting a comprehensive source of drug repurposing opportunities when the two approaches are combined.

## Introduction

Immune-mediated diseases (IMDs) arising from dysregulated immune responses affect up to 10% of the global population, posing a major health burden[1,2]. Both innate and adaptive arms of immunity contribute to pathogenesis of IMD via altered immune cell frequencies and activation states[3]. Dysfunctional immune cells, particularly T cells and B lymphocytes, are central to the pathogenesis of these disorders, leading to the production of inflammatory mediators like cytokines and autoantibodies.

Despite the substantial burden of autoimmune and inflammatory diseases, the pharmaceutical arsenal remains limited, owing to the complexity of the dysfunctional immune cascade underlying these heterogeneous conditions[4,5]. The absence of disease-specific biomarkers and need for chronic therapy compound the challenges of developing targeted agents. Therefore, given shared immune pathogenic pathways across IMD, there is substantial interest in repurposing of drugs in this disease category. Drug repurposing involves demonstrating the efficacy of a drug, previously tested for safety and effectiveness in one medical condition, for a different indication[6]. For example, adalimumab, an anti-tumour necrosis factor (TNF) monoclonal antibody has been initially approved for treatment of rheumatoid arthritis but has since been extended for use in psoriasis, ankylosing spondylitis, Crohn’s disease, and ulcerative colitis[7].

Since mid-2000s, genome-wide association studies (GWAS) have uncovered hundreds of disease-associated loci implicating immune-related genes[8]. Uncovering specific proteins driving IMD holds promise for new targeted therapeutics. Human genetic support can more than double the approval odds of drug target in preclinical development, as well as progression along subsequent phases of clinical trials[9,10]. In general, the strongest support is provided by variants with impact on protein-coding sequence of gene as they offer the least ambiguous mapping to a drug target, unlike intergenic and intronic variants[11].

Mendelian randomization (MR) is one approach that can help prioritise targets in the drug development pipeline by explicitly modelling causal relationships. MR uses GWAS-derived genetic variants as instrumental variables (IV) to study the lifetime effects of genetic perturbations of drug targets. This allows for the examination of causal effects on a chosen outcome of interest, such as any of immune-mediated diseases[12]. By leveraging the natural random assortment of genetic material during meiosis, MR provides a powerful framework to assess causality, mitigating issues of reverse causation and confounding that often plague observational studies[13].

For reliable causal inference within the MR framework, certain essential assumptions must be met. These include the requirement that the genetic variants employed as proxies exhibit a robust association with the targeted exposure (known as the *relevance* assumption). Additionally, it is crucial that the relationships between IV genetic variants and both the exposure and the outcome are not influenced by confounding factors such as environmental variables, genetic ancestry or co-inherited genetic signals (referred to as the *independence* or *exchangeability* assumption). Moreover, it is necessary to ensure that the genetic variants’ association with the outcome is not influenced by pathways independent of the primary exposure, a phenomenon known as horizontal pleiotropy (*exclusion restriction* assumption).

We can identify two main approaches in MR analyses looking at causal support for a given drug target against a disease indication. While the outcome GWAS used involve primarily disease incidence, the exposure GWAS can instrument a direct measure of gene expression (either messenger RNA or protein) in disease-relevant tissue or a downstream biomarker or clinical risk factor (here referred to as “intermediate trait”)[14]. In the first approach, variants related to two molecular phenotypes: protein and mRNA abundance are known as protein and expression quantitative trait loci (QTL), respectively. Typically, protein QTL are preferable given the closer relationship of protein levels to clinical phenotypes and mechanism of drug action which usually targets proteins[15]. An important constraint in the analytical application of pQTLs is the potential for confounding arising from artefactual effects caused by differential capture affinity driven by protein-altering variants (PAV) rather than biologically meaningful changes in protein concentration[16]. To address these biases, it is recommended to explore multiple independent pQTL datasets, incorporating different assays for the measurement of protein levels whenever possible[17].

In the intermediate trait MR, exposure variants are extracted from GWAS for relevant disease risk factors or biomarkers with available lead variant(s) located in the drug target gene of interest. This approach offers confirmation that the genetic variant indeed influences the clinical outcome of interest[14]. Previously employed intermediate traits in MR include HbA1c for proxying effects of GLP-1 agonists[18], a class of antidiabetic medication, CRP for mimicking the effect of IL-6 signalling[19] and height for the effect of NPR2 and NPR3 signalling on cardiovascular disease[20]. However, limited use has been made of this approach in evaluating drug targets for IMD. Here, we propose a new intermediate trait category for use in proxying drug targets in IMD: immune cell abundance. There is substantial animal model, observational and MR evidence[21–26] implicating immune cell dysregulation in the pathogenesis of immune-mediated disorders, ranging from roles of T_H_2 cells, eosinophils, and neutrophils in allergic conditions like asthma[27,28] and eczema[29,30] to expansion of certain T helper lymphocyte and B lymphocyte lineages in autoimmune diseases such as systemic lupus erythematosus (SLE)[31], multiple sclerosis[32] and rheumatoid arthritis[5]. Furthermore, approved drug targets for IMD are usually classed as immunosuppressants or immunomodulators, which alter the balance of immune blood cells in their course of on-target action. For example, corticosteroids reduce the number of various immune cells, including lymphocytes and eosinophils in the blood[33], while methotrexate promotes monocyte apoptosis[34] and mycophenolate mofetil suppresses T and B lymphocyte proliferation[35].

Here, we implement a multi-pronged MR strategy exploiting genetic instruments from 23 large-scale GWAS resources. Firstly, we examine causal connections between blood cell immunotypes and 14 IMDs. Composition of specialized white cells in peripheral blood underpins immunocompetence, making these genetics-derived perturbations clinically relevant. We present a comprehensive examination of bidirectional relationships between immune cell counts and IMD.

In the next stage of our MR analyses, we prioritize genes as potential therapeutic targets for IMD using intragenic variants from immune cell GWAS established as causal, using a novel intermediate trait category in MR studies of autoimmune and inflammatory disease. As proteins constitute ultimate drug-actionable targets, we also employ an alternative approach leveraging blood serum protein QTLs (pQTLs) to instrument targets. To check the robustness of our findings, we include pQTLs derived in 4 largest independent studies so far, which use two different technical assays. Lastly, we confirm all our MR findings with the colocalisation sensitivity analysis.

By evaluating concordance and complementarity of these MR approaches, we aim to validate known and discover novel drug target indications for immune-mediated disease. Integrative evaluation of previously implicated genes and their repurposing opportunities using existing drugs can accelerate translation of genetic insights to the clinic.

## Materials and Methods

### Immune blood cell exposures

We gathered a comprehensive selection of count-based peripheral immune cell GWAS (**Supplementary Table 1**) derived from complete blood count, which included counts of basophils, eosinophils, granulocytes, lymphocytes, monocytes, neutrophils, myeloid white cells, total white blood cells and also their relative percentages. The GWAS were conducted using mostly the UK Biobank data of 132,959-456,785 European ancestry individuals[36–38].

### Immune-mediated disease outcomes

We compiled a selection of 14 IMD GWAS (**Supplementary Table 2**): ankylosing spondylitis (AS)[39], asthma[40,41], chronic sinusitis[39], eczema (atopic dermatitis)[39,42], eosinophilic esophagitis[43], psoriasis[44], juvenile idiopathic arthritis[45], rheumatoid arthritis[46], inflammatory bowel disease (IBD)[47], Crohn’s disease[47], ulcerative colitis[47], multiple sclerosis (MS)[48], systemic lupus erythematosus[49], type 1 diabetes[50]. We employed European-ancestry GWAS with the highest power (as judged by sample size and number of top loci) whenever available. The biggest GWAS for two IMD: asthma[41] and eczema[42] included a significant contribution from UK Biobank (N > 300,000). As two-sample MR estimates can be biased by population sample overlap between exposure and outcome GWAS[51], for these traits we included replicate, independent GWAS[39,40] to confirm that the relationships detected in the two-sample MR analyses involving the immune cell exposures and Olink pQTLs (*see below*, also derived from UK Biobank).

### Protein quantitative trait loci (QTL) exposures

Blood plasma pQTL data was obtained from large European-ancestry studies with protein abundance measured using two different affinity-based technologies (**Supplementary Table 3**): aptamer-based SomaScan ver 4 including ∼4,500 protein targets (ARIC[52], deCODE[53], FENLAND[54]) and antibody-based Olink assay with ∼3,000 targets (UK Biobank[55]).

### Selection of genetic instruments

To identify genetic instruments for each exposure, we selected SNPs demonstrating a robust association at the genome-wide significance threshold (*p*-value < 5 × 10^-8^). Subsequently, we conducted clumping of these SNPs to ensure that the linkage disequilibrium (LD), measured by r^2^, was maintained at less than 0.001 within a 10 Mbp range in the 1000 Genomes European panel[56]. This step aimed to prevent multiple instruments from capturing the same causal effect and assure their independence. The clumping process was executed using plink version 1.943[57], facilitated by the ld_clump function in the ieugwasr R package (available at https://mrcieu.github.io/ieugwasr). In each MR analysis, we extracted and harmonized genetic variant associations for the outcome trait. Subsequently, mean F-statistics and R^2^ were computed to assess potential weak instrument bias.

For intermediate trait instruments, prior to clumping, we obtained all genome-wide significant (*p*-value < 5 x 10^-8^) hits overlapping genes in the immune cell GWAS showing a robust, high-confidence association with immune-mediated disease. Using Variant Effect Predictor (VEP)[58] annotations, we subsequently selected the variants with the highest priority annotation (reflecting intragenic location and expected severity of functional consequence) available for each gene (**Supplementary Table 4**).

For pQTL exposures we included the following additional steps due to nature of molecular phenotypes resulting in high risk of bias from horizontal pleiotropy[59]. First of all, we only used *cis*-pQTLs, defined as positioned maximum 1 Mbp away from gene’s TSS and discarded trans-pQTLs as *cis-* variants are less likely to be affecting the outcome through expression of multiple genes [60]. Secondly, we flagged SNPs (or correlated proxies at r^2^ > 0.6) with potentially epitope-altering mutations (such as: “stop_gained", "stop_lost", "frameshift_variant", "start_lost", "inframe_insertion", "inframe_deletion", "missense_variant", "protein_altering_variant”) which could result in artefactual variation in protein levels. However, we did not remove them due to their high frequency in the exposure datasets and minority of them (∼25%) having been estimated to result in false positive pQTLs[61].

### Two-sample Mendelian Randomization

Two sample MR analyses (Wald ratio for single-SNP instruments or inverse variance weighted regression, IVW, for multi-SNP instruments) were carried out using TwoSampleMR R package[62]. For instances where the instrument comprised three or more SNPs, we conducted sensitivity analyses incorporating MR-PRESSO which can identify and adjust for pleiotropic outlier variants[63], weighted median, weighted mode, and MR-Egger methods to ensure the consistency of estimates. Additionally, we computed I^2^ and Cochran’s Q to investigate the variability of estimates among the variants included in each instrument. The MR-Egger intercept test was applied to assess the potential impact of directional pleiotropy on our results[64]. To evaluate the NOME assumption for MR-Egger, we computed the I^2^_GX_ statistic as an indicator of potential attenuation bias [65].

In the case of all MR analyses evaluating the effect of immune cells on IMD, where we had strong prior evidence of reverse relationships in which IMD may effect immune cell abundance[5], we conducted bidirectional MR. In addition, to examine whether the potential causal effects were independent of the outcome influencing the exposure, we also included Steiger filtering[30] in all our analyses.

### Colocalisation

We employed the Bayesian colocalisation method coloc[66] to assess whether the nominally significant (p-value < 0.05) MR effects observed for drug targets were indicative of causation or potentially confounded by linkage disequilibrium (LD)[67]. Among the pQTL-instrumented MR analyses, the only three studies (ARIC, deCODE, UK Biobank) with published full summary statistics were used to run coloc. A 100 kb window around each SNP in the instrumental variable was used to define the colocalisation region and analysis was conducted if at least 50 variants were identified in the specified window.

### Drug target validation and repurposing

Protein target-disease indication pairs for drugs with approved (globally), in active development (preclinical and in trials) and ceased (no development recorded for > 1 year) status were downloaded from manually curated Pharmaprojects database (https://www.citeline.com/en/products-services/clinical/pharmaprojects), separately for every IMD included, on 29^th^ August 2023 (**Supplementary Table 14**). Pharmaprojects has been a continuously updated industry-standard reference for pharmaceutical industry for over 40 years[68]. In cases of multiple drugs targeting the same protein, we selected the drugs with the most advanced development status. In addition, a manually curated list of 4,723 genes involved in immune response derived from the ImmPort[69] project was accessed via InnateDB[70].

## Results

### Bidirectional causal effect of immune blood cell counts on IMD

We first wanted to establish robust causal associations between peripheral immune cells and immune-mediated disease using Mendelian Randomization (**Supplementary Figure 1**). We ran a number of sensitivity MR analyses, in addition to the baseline IVW analyses involving all genetic instruments (**Supplementary Table 5**). IVW was also run excluding the major histocompatibility complex (MHC) region located at chromosome 6, between 28.5Mb–33.5Mb. This region is prone to skewed results due to complex LD patterns resulting in potential for including multiple correlated variants[71]. In addition, we also conducted outlier-adjusted MR-PRESSO test including and excluding the MHC region (**Supplementary Table 9**). The nominally significant results (p-value < 0.05) across the 4 analyses were intersected to produce a high-confidence set of 59 immune cell phenotype->IMD associations (**Supplementary Table 8**). For two IMD outcomes (asthma and eczema) we included an independent replicate GWAS due to UK Biobank-derived sample overlap between exposure and outcome GWAS when using the largest available IMD GWAS. Across the duplicate outcome GWAS, we found a directionally robust pattern of associations, regardless of sample overlap presence (**Figure 1A**) so in the subsequent analyses we focussed on results derived from the biggest, single representative GWAS for each disease.

**Figure 1.**
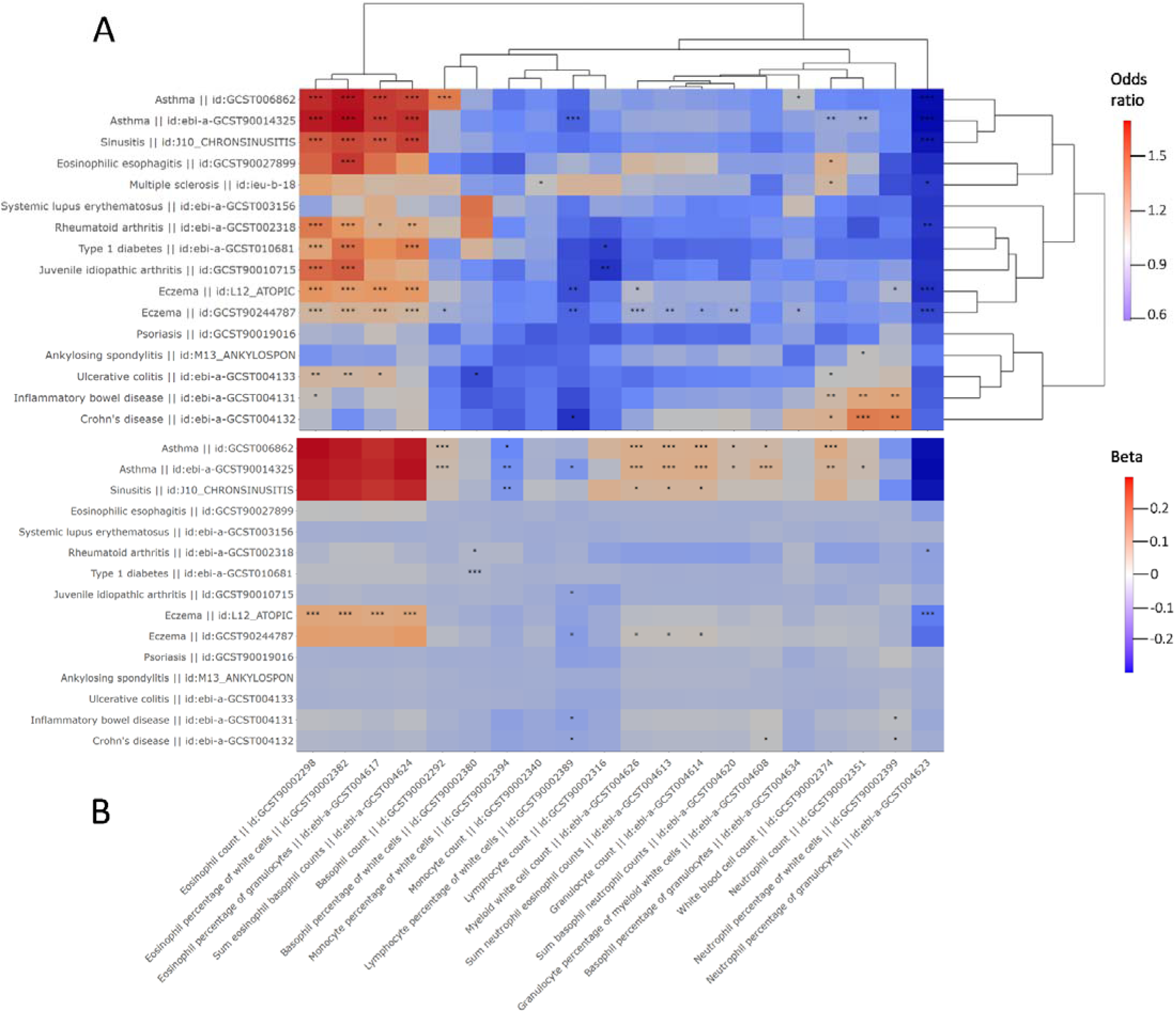
**A**) A complete-linkage clustered matrix of the central odds-ratio estimates of the associations between peripheral immune cells (x-axis) and immune-mediated disease (IMD y-axis). High-confidence associations with p-values of: < 0.05 (*), <0.005 (**), <0.0005 (***) in the IVW analysis excluding the MHC region are highlighted with an asterisk(s). **B**) Heatmap of the central beta estimates of the associations between immune-mediated disease (IMD, y-axis) and immune cell counts (x-axis). High-confidence associations with p-values of: < 0.05 (*), <0.005 (**), <0.0005 (***) in the IVW analysis excluding the MHC region are highlighted with an asterisk(s). Multiple sclerosis is missing from the heatmap as no instrument outside of the MHC region was available for this exposure.

Overall, we discovered a strong positive effect of eosinophil phenotypes (count, percentage of white cells, percentage of granulocytes, sum eosinophil basophil counts) on genetic susceptibility to multiple disease (**Figure 1A**). These included disease with atopy component: asthma (OR=1.72, CI_95%_=1.6-1.85, p-value=3.9×10^-46^), eczema (OR=1.25, CI_95%_=1.17-1.33, p-value=1.7×10^-11^), sinusitis (OR=1.57, CI_95%_=1.45-1.69, p-value=5.1×10^-31^) and eosinophilic esophagitis (OR=1.6, CI_95%_=1.23-2.1, p-value=4.8×10^-4^), but also rheumatoid arthritis (OR=1.36, CI_95%_=1.18-1.58, p-value=3.7×10^-5^), juvenile idiopathic arthritis (OR=1.51, CI_95%_=1.24-1.84, p-value=3.7×10^-5^), type 1 diabetes (OR=1.47, CI_95%_=1.26-1.72, p-value=1.6×10^-6^) and ulcerative colitis (OR=1.22, CI_95%_=1.06-1.4, p-value=5×10^-3^). The IVW estimates given correspond to 1 SD change in the exposure, here eosinophil percentage of white cells.

Using MR, we also assessed the reverse causal pathway: from IMD to immune cell counts (**Figure 1B**). There, we found some evidence that the relationship between eczema, asthma, sinusitis and eosinophil phenotypes is bidirectional. One unit increase in the log odds of eczema was associated with increase of eosinophil percentage of white cells (β=0.103, CI_95%_=0.058-0.149, p-value=8×10^-6^). For asthma and sinusitis, that direction of relationship was not robustly confirmed when Steiger filtering was applied (**Supplementary Table 13**), which suggested that the main causal direction was from eosinophils to asthma and sinusitis.

We found a positive relationship between neutrophil counts (also neutrophils percentage of white cells) and bowel disease: IBD (OR=1.29, CI_95%_=1.11-1.51, p-value=9.4×10^-4^) and Crohn’s disease (OR=1.44, CI_95%_=1.19-1.74, p-value=1.8×10^-4^). On the other hand, we found a negative causal relationship between neutrophil percentage of granulocytes and atopic disease (asthma: OR=0.62, CI_95%_=0.55-0.71, p-value=6.4×10^-13^, eczema: OR=0.8, CI_95%_=0.73-0.88, p-value=8.8×10^-6^, sinusitis: OR=0.65, CI_95%_=0.58-0.74, p-value=1.8×10^-12^), as well as rheumatoid arthritis (OR=0.76, CI_95%_=0.64-0.89, p-value=1.1×10^-3^). Similar to eosinophils, bidirectional relationship was only robustly supported for eczema (β=-0.088, CI_95%_=[-0.132, - 0.044], p-value=7.5×10^-5^), but not asthma and sinusitis.

Negative effect of immune cell type abundance on IMD was also found for lymphocytes. Lymphocyte percentage of white cells was negatively associated with the odds of asthma (OR=0.89, CI_95%_=0.83-0.95, p-value=4.5×10^-4^), eczema (OR=0.83, CI_95%_=0.75-0.93, p-value=7.9×10^-4^) and Crohn’s disease (OR=0.74, CI_95%_=0.59-0.93, p-value=0.01), while lymphocyte count had a negative association with genetic susceptibility to type 1 diabetes (OR=0.78, CI_95%_=0.65-0.94, p-value=7.3×10^-3^) and juvenile idiopathic arthritis (OR=0.74, CI_95%_=0.61-0.91, p-value=4.4×10^-3^). In addition, we found some evidence (p-values: 0.01 – 0.04) for bidirectional relationship between lymphocyte percentage of white cells with five IMD (**Figure 1B**).

Genetic liability to asthma and sinusitis were revealed as a potential risk factor across a number of immune cell phenotypes (**Figure 1 B**). We found a potential negative causal relationship between asthma (β=-0.055, CI_95%_=[-0.086, -0.024], p-value=5.9×10^-4^), sinusitis (β=-0.042, CI_95%_=[-0.067, -0.018], p-value=6.3×10^-4^) and monocyte percentage of white cells but the relationship was not replicated using the absolute monocyte counts outcome.

Intriguingly, we found no significant genetic support for causal association between any immune cell phenotypes and psoriasis as well as systemic lupus erythematosus (**Figure 1 A**). For ankylosing spondylitis and multiple sclerosis, we only found fairly weak statistically significant support (p-value = 0.005-0.05) for immune cell basis of susceptibility (AS - neutrophil count, MS - monocyte count, white blood cell count, and neutrophil percentage of granulocytes).

As expected due to genetic complexity of exposure and outcome phenotypes, sensitivity analyses revealed a high amount of heterogeneity[72] in the MR IVW estimates using Cochrane’s Q (**Supplementary Table 6, 11**) but not horizontal pleiotropy as measured by MR-Egger intercept test (**Supplementary Table 7, 12**)

### Drug target prioritisation using intermediate trait MR

Having established immune blood cells are putatively causally associated with immune-mediated disease, we were then able to prioritise drug targets using intermediate trait MR. In this analysis, we utilised genic variants robustly associated with immune cell abundance (p-value < 5 x 10^-8^) as proxy instruments. In total, we were able to instrument 1,081 variants in 371 genes (**Supplementary Table 16**) out of 834 previously targeted for treatment of any of 12 immune-mediated disease (**Supplementary Table 14**). A total of 6,127 MR analyses were conducted, with some genes possessing instruments across a number of intermediate traits and so analysed independently (**Supplementary Table 15).** A quarter of IVW/Wald MR results (1,728) displayed evidence of association at the nominal significance level (p-value < 0.05), which revealed 811 unique gene-IMD associations. These involved 261 genes with the highest number of hits obtained for asthma (121 genes), inflammatory bowel disease (109 genes), eczema (83 genes) and chronic sinusitis (54 genes). Overall, we found extensive overlap of majority of drug targets with nominally significant MR evidence across IMD (**Supplementary Figure 2**). The majority of genes with MR evidence (193, 74%) are known to contribute to immune function, which supports the choice of intermediate immune cell phenotypes.

Since MR results are liable to confounding by linkage disequilibrium on their own, we also sought confirmation with the colocalisation approach, which assess the probability of shared genetic signal between the exposure and the outcome (**Supplementary Table 20**). **Figure 2** highlights the MR results for genes with strong colocalisation support (PP_H4_ > 0.8) for above-mentioned 4 IMD, using instruments sourced from eosinophil count GWAS: **A**) asthma – 27 genes, **B**) inflammatory bowel disease – 12 genes, **C**) sinusitis – 11 genes, **D**) eczema – 10 genes. None of the targets were common to all the 4 conditions, but *IL3* and *GATA3* were shared among IBD, sinusitis and asthma, *CDK2*, *IL1R1* among eczema, asthma and sinusitis, *IL33* and *IL7R* between asthma and sinusitis, *ERBB3*, *PRKCQ*, *STAT6* between asthma and eczema, *CSF2*, *FADS1* and *FAP* between IBD and asthma, *PPARG* between sinusitis and eczema, *GPR65* and *IL2* between eczema and IBD, and finally *PTPN11* was shared between IBD and sinusitis. Many of these are not just confirmatory for existing indications for approved drugs or drugs in development (*PRKCQ*, *IL1R1, STAT6, IL2*) but also suggesting additional repurposing opportunities for other IMD (*IL7R*, *CDK2, GATA3, IL33, GPR65, PTPN11, ERBB3, FAP*). However, opposite direction of effect across IMD was found for four targets (*IL3*, *CSF2*, *FADS1*, *PPARG*) indicating lack of feasibility for drug repurposing and even potential adverse side effects.

**Figure 2.**
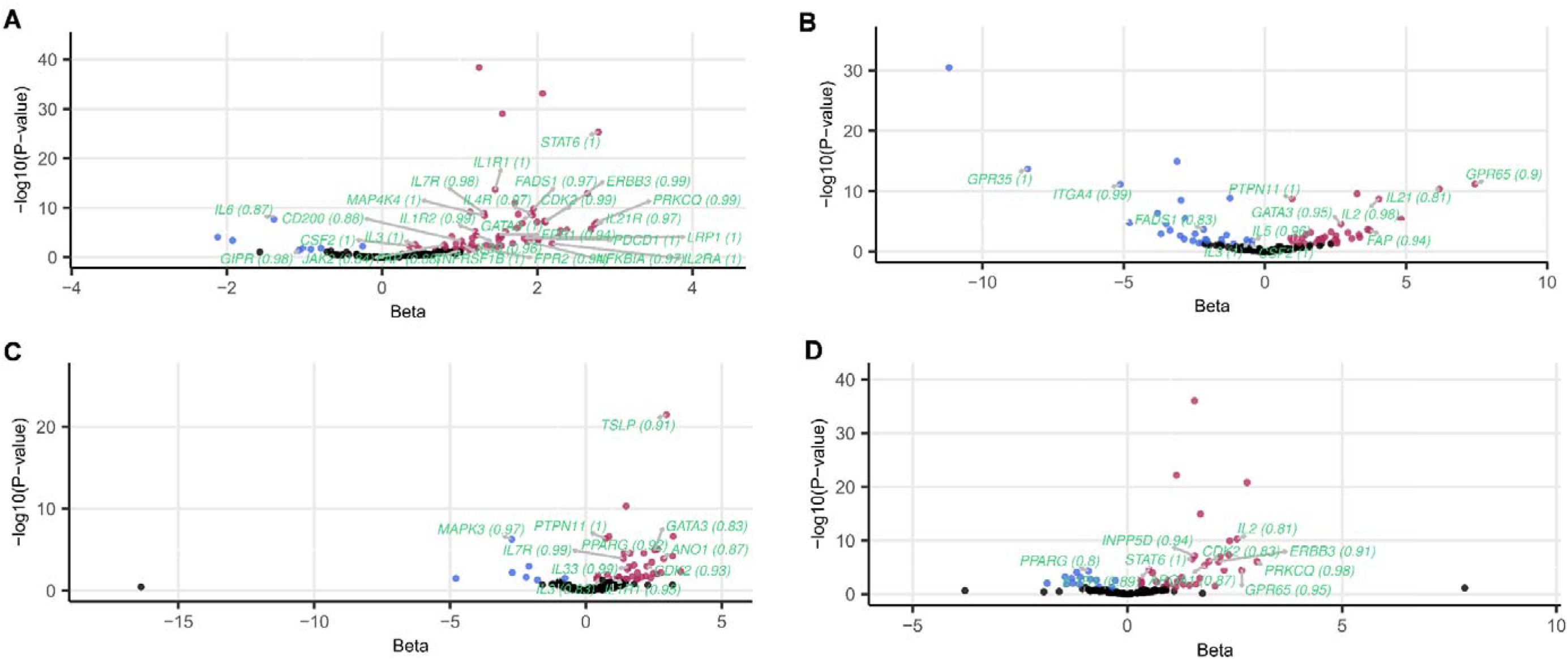
Intermediate trait MR results for 4 IMD showing the highest number of hits with strong colocalisation support (PP_H4_ ≥ 0.8): **A**) asthma, **B**) inflammatory bowel disease, **C**) chronic sinusitis, **D**) eczema. The x-axis represents the central effect size estimate and y-axis represents the -log10(P-value) in MR analysis, while the gene symbols and colocalisation probabilities of nominally significant MR hits (p-value < 0.05; beta > 0 in red, beta < 0 in blue) with strong colocalisation evidence highlighted in light green andPP_H4_ values listed in parenthesis. We used eosinophil counts as GWAS source for exposure instruments.

Overall, we found 339 MR results at nominally significant level which were supported by strong colocalisation signal. Among 169 unique gene-IMD associations with robust coloc evidence, we found that a minority of associations (62, 36.7%) were confirmatory of existing drug target indications and the majority, 107 (63.3%), were novel.

We also provide additional MR sensitivity statistics for heterogeneity (**Supplementary Table 17**), pleiotropy (**Supplementary Table 18**) and Steiger filtering (**Supplementary Table 19**). Since heterogeneity and pleiotropy tests rely on multi-SNP instruments (≥ 2 and ≥ 3 SNPs, respectively), we only obtained their results for a limited number of MR analyses (392 and 45, respectively). High levels of estimate heterogeneity (p-val < 0.05) were only seen for 86 out of 392 associations (22%) and no significant evidence for pleiotropy was found. The main direction of causal effect from immune cell-instrumented drug target to IMD was confirmed for the majority of MR associations (5,352, 87.3%) using Steiger filtering.

### Drug target prioritisation using protein QTL MR

Next, we leveraged an alternative source of instrumental variables for drug targets for IMD: protein QTL corresponding to genetic associations with circulating protein concentration in the blood serum. For comparison purposes, we used three studies which applied the SomaScan technology for protein measurement (ARIC, deCODE and FENLAND), and one study which used the Olink technology (UKBioBank, UKBB). Across the four cohorts combined, we were able to obtain strong instruments for 361 proteins (233, 193, 169, 280 in ARIC, deCODE, FENLAND and UKBB, respectively, **Supplementary Table 23**) out of 834 with IMD indication in Pharmaprojects, which were used to run 11,370 cohort-specific MR analyses (**Supplementary Table 22**). Over 10% of IVW/Wald MR results (1,431) showed evidence of association at the nominal significance level (p-value < 0.05), which reflected 841 distinct gene-IMD associations. These included 284 unique proteins with the highest number of hits returned for inflammatory bowel disease (82 proteins), Crohn’s disease (80 proteins), rheumatoid arthritis (75 proteins) and psoriasis (72 proteins). Similar to intermediate trait MR, we uncovered ubiquitous sharing of drug targets with nominally significant MR evidence across IMD (**Supplementary Figure 3**) and a high proportion of targets (78.7%) with immune-related function.

Comparison of MR results obtained from the four cohorts using two dimensionality reduction methods (hierarchical clustering-**Supplementary Figure 4** and principal component analysis -**Supplementary Figure 5**) showed a strong effect of a particular protein measurement chemistry. SomaScan-sourced pQTLs (ARIC, deCODE, FENLAND) clustered distinctly away from Olink (UKBB), underscoring the importance of inclusion of instruments from diverse sources. When considering overlap of gene-IMD nominally significant associations across all the 4 pQTL sources (**Supplementary Figure 6 A**), the top 3 categories contained singletons present only in UKBB (273 associations), ARIC (127 associations), deCODE (83 associations), followed by 63 associations shared across the 4 cohorts. However, this result is mostly driven by *a priori* limited sharing of genetic instruments across cohorts and when considering MR analyses involving only shared drug targets, the top category is composed of significant MR hits across all the four cohorts (**Supplementary Figure 6 B**). Analogous conclusions can be drawn when sub-setting to gene-IMD associations significant at stringent Bonferroni-corrected p-value threshold (p-value < 10^-7^, **Supplementary Figure 6 C-D**).

Confirmation of MR results with colocalisation (**Supplementary Table 27**) revealed strong support (PP_H4_ ≥ 0.8) for 230 MR associations (16% out of 1,431) comprised of 83 distinct protein-IMD associations. In keeping with results from intermediate trait MR, two-thirds of these associations were novel (53) rather than confirmatory of existing disease indications for drug target (30). **Figure 3** highlights the advantage of using multiple pQTL sources (**A** – ARIC, **B** – deCODE, **C**-UK Biobank) for ulcerative colitis, the IMD showing the highest number of hits with strong colocalisation support. *VSIR* and *CD274* loci robustly colocalised with pQTLs across the 3 cohorts, while evidence for colocalisation for *STAT3* was found both in ARIC and deCODE. Additionally, UKBB pQTLs independently prioritised 6 proteins (*CD6*, *IL10*, *IL10RA*, *IL1RL1*, *ITGAV*, *OSMR*), none of which were instrumented in the other 3 pQTL studies.

**Figure 3.**
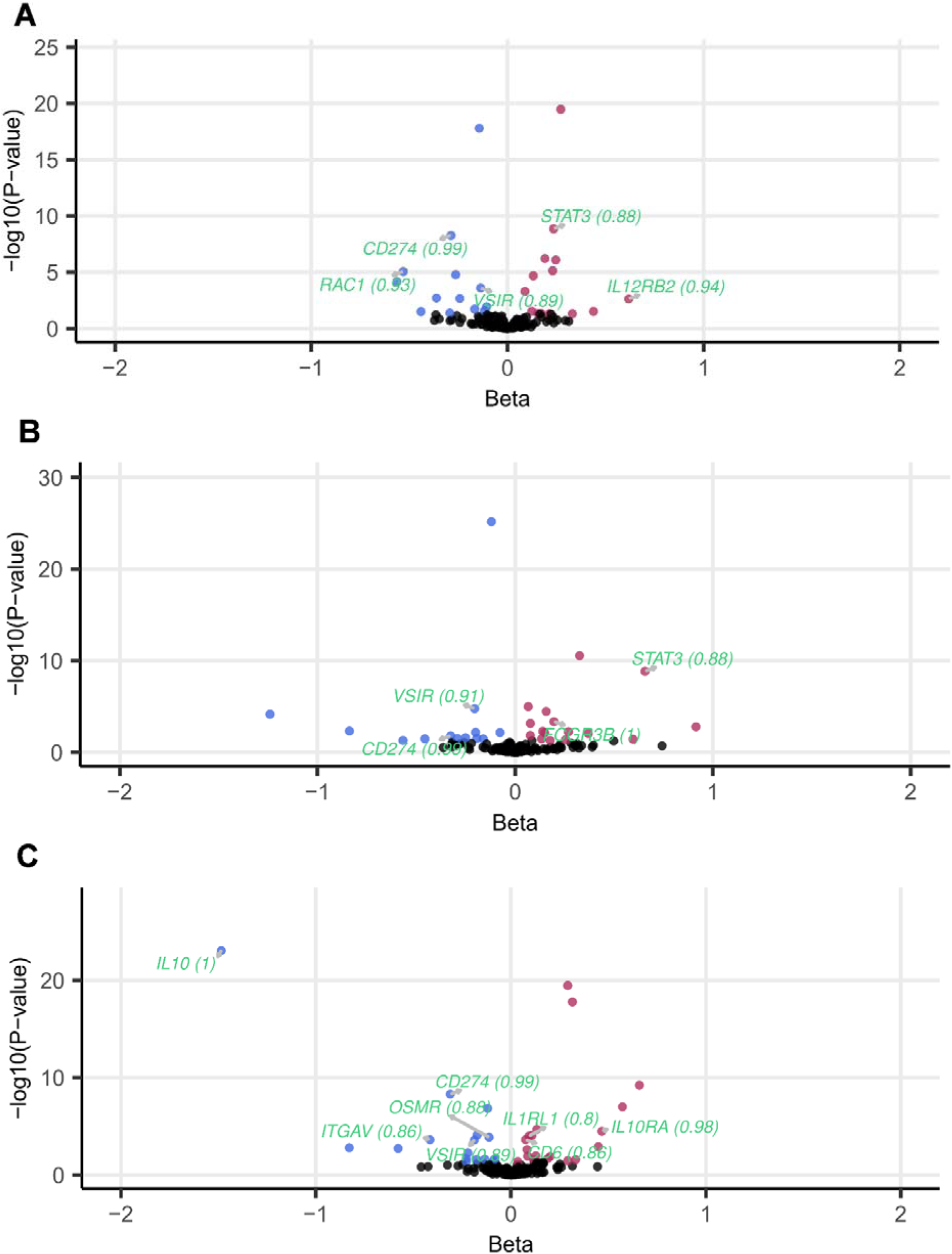
pQTL MR results for ulcerative colitis, the IMD showing the highest number of hits with strong colocalisation support (PP_H4_ ≥ 0.8). Plot compares MR results across different blood plasma pQTL exposure sources: **A**) ARIC, **B**) deCODE, **C**) UK Biobank. The x-axis represents the central beta estimate and y-axis represents the - log10(P-value) in MR analysis, while the gene symbols and colocalisation probabilities of nominally significant MR hits (p-value < 0.05; beta > 0 in red, beta < 0 in blue) with strong colocalisation evidence highlighted in light green and PP_H4_ values listed in parenthesis.

Among the MR analyses with sufficient number of exposure SNPs for heterogeneity analysis (**Supplementary Table 24**), only a small number showed significant evidence (p-value < 0.05) against the null hypothesis of homogeneity (297 out of 2875, 10%). MR Egger intercept test (**Supplementary Table 25**) revealed only 8 MR results (out of 1,576) with evidence for pleiotropy, 4 of which involved *IL5RA* pQTL used as exposure. Using Steiger filtering (**Supplementary Table 26**), the direction of causal effect from protein QTL to IMD was confirmed for all the MR associations, bar 3 for the *BRD2* protein.

### Integrative MR-based validation and repositioning of drug targets

We used Pharmaproject’s list containing approved drug targets to disease indication assignments to compare the sensitivity of intermediate trait MR (**Supplementary Table 21**) and protein QTL MR (**Supplementary Table 28**) approaches. Sensitivity was generally poor but with a large variance across disease outcomes. Mean intermediate trait MR sensitivity was estimated at 0.49 (± 0.23 SD), while for pQTL MR the mean sensitivity was 0.28 (±0.12 SD); similar results were obtained when focussing on drug targets in development.

Given fairly low predictive power of individual MR approaches, we were interested to compare how much additional information combining them can provide. Intersecting all nominally significant gene-IMD associations found in intermediate trait and pQTL MR (**Figure 4 A**) showed only 13% overlap (190 associations). The remainder of associations were symmetrically distributed between intermediate trait MR (621, 42%) and pQTL MR (651, 45%). When sub-setting to MR analyses which instrumented the same genes across both approaches (**Figure 4 B**), the fraction of overlap substantially increased (39%), with slightly more associations found only in intermediate trait MR (177, 36%) than pQTL MR (122, 25%). Altogether, we counted 167 highest confidence MR hits with p-value < 10^-7^ (Bonferroni-corrected threshold), with 77 (46%) specific to intermediate trait MR, 60 (36%) shared, and 30 (18%) specific to pQTL MR (**Figure 4 C**). The vast majority of singleton associations were due to availability of a given gene exposure only through one approach. Only 9 and 2 gene-IMD associations were found solely via intermediate trait MR and pQTL MR, respectively (**Figure 4 D**), when limiting the analysis to the subset of MR results with overlapping target gene exposures.

**Figure 4.**
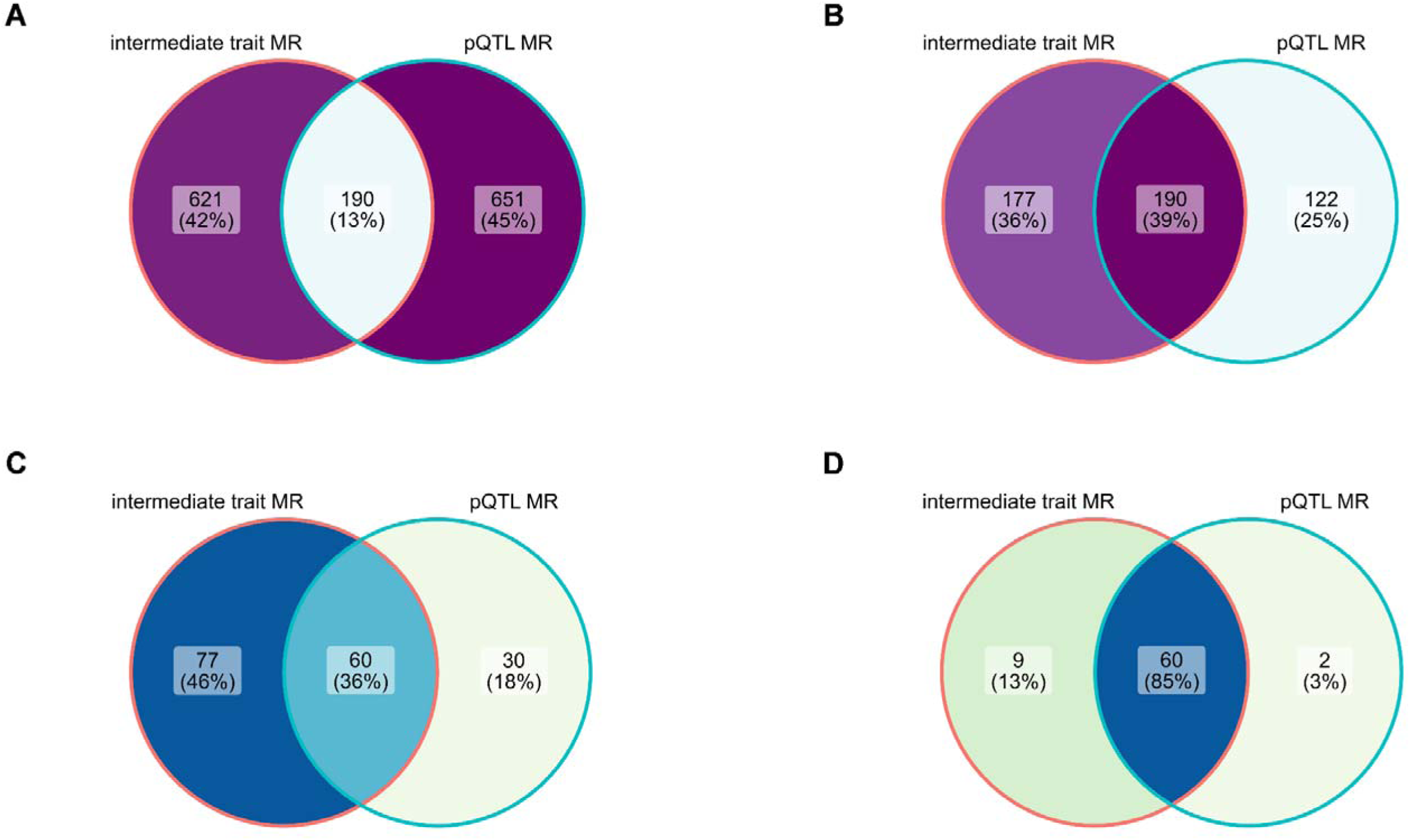
Venn diagram showing gene-IMD association overlap between significant intermediate trait MR and pQTL MR results. **A**) nominal p-value threshold met, including all tested genes with a suitable instrument; **B**) nominal p-value threshold met, including only overlapping target genes between intermediate and pQTL instrument sources; **C**) Bonferroni-corrected p-value threshold (p-value < 10^-7^)* met in either set, including all tested genes with a suitable instrument; **D**) Bonferroni-corrected p-value threshold met in either set, including only overlapping target genes between intermediate and pQTL instrument sources. *Bonferroni-corrected threshold equalled: 0.05 / (number of proteins tested x number of outcomes tested)

We summarised information regarding the drug targets with the highest confidence evidence present in both intermediate trait MR and pQTL MR in **Table 1**. We found genetic evidence for repurposing opportunity in asthma and eczema for already approved rheumatoid arthritis drug anakinra which targets *interleukin 1 receptor, type I* (*IL1R1*). For pateclizumab, an antibody directed towards *lymphotoxin alpha* (*LT*α) which had been halted for development as a rheumatoid arthritis medication due to lack of efficacy[73], we found genetic evidence for potential in treatment of 3 other IMDs. In addition, we discovered strong MR support for *STAT6* inhibition in asthma, currently in preclinical development by Recludix and Sanofi.

For drugs in phase 1 clinical trials, we identified potential repurposing opportunities across 2 other IMD for a psoriasis target *bromodomain-containing 2* (*BRD2)* kinase. Interestingly, validation of disease indication was found for *CD200 receptor 1* (*CD200R1*) and eczema, as well as *tumor necrosis factor receptor 1* (*TNFRSF1A*) and multiple sclerosis but misaligned direction of effect suggests that the drugs currently being trialled could potentially fail. For drug targets in phase 2 trials, we found confirmatory MR evidence for *interleukin-7 receptor* (*IL7R*) and eczema, multiple sclerosis, as well as *STAT3* and Crohn’s disease. On the other hand, repurposing opportunities were uncovered for *interleukin 4* (*IL4*), *IL7R* and asthma, *STAT3* and ulcerative colitis.

Among drugs in the phase 3 clinical trials, we found the highest number of repurposing opportunities (4 IMD) for azeliragon, a small-molecule inhibitor of *AGER*. We also provide genetic validation for cytokine targets of monoclonal antibodies: *interleukin 23 receptor* (*IL23R*) for IBD with Crohn’s disease (but featuring misaligned direction of effect relative to currently developed drug), as well as *colony stimulating factor* 2 (*CSF2*), *interleukin 1 receptor like 1* (*IL1RL1*), *interleukin 33* (*IL33*) for asthma. For *CSF2* and *IL1RL1* inhibitors, MR suggests eczema as an additional disease indication, while for *IL1RL1* and *Fc* γ *receptor IIb* (*FCGR2B*) MR delivered repositioning evidence favouring inflammatory bowel disease.

## Discussion

This study leveraged the Mendelian Randomization method to establish genetically-predicted immune cell mediation of IMDs. We then conducted MR analyses involving immune cell abundance-derived and protein QTL-derived genetic instruments to systematically prioritize indications for approved and in-development IMD drug targets, highlighting avenues for repurposing and providing validation for drugs in preclinical stages and in clinical trials. Integrating multiple MR approaches with colocalisation strengthens confidence in our findings, while indicating that a substantial proportion of MR results may stem from LD patterns between distinct causal variants rather than true causality.

Using a crude measure of immune system activation – immune cell abundance (absolute and relative), we sought to establish bidirectional causal relationships of broad immune cell categories with immune-mediated diseases. Notably, eosinophil phenotypes emerged as key players, exhibiting a strong positive effect on diseases with atopic components. The observed bidirectional causal effect between eosinophil phenotypes and specific diseases aligns with prior research implicating eosinophils in the pathogenesis of atopic conditions such as asthma, eczema and eosinophilic esophagitis with their mechanistic contribution to type 2 inflammation[74,75]. Tissue eosinophilia is linked to epithelium remodelling along with airway hyperreactivity manifestations in asthma as well as skin and oesophageal mucosa infiltration in eczema and eosinophilic esophagitis, respectively. Reverse positive association of asthma, eczema and sinusitis with eosinophils could be attributed to secondary immunologic activation resulting in positive feedback loop in the atopic march[76,77]. Less expected was a robust MR relationship of eosinophiles with a range of autoimmune diseases: rheumatoid arthritis, juvenile idiopathic arthritis, type 1 diabetes and ulcerative colitis, which requires further investigation and triangulation.

We found a strong positive association between genetically predicted neutrophil counts and incidence of inflammatory bowel disease and Crohn’s disease. In observational studies of IBD initiation, neutrophils have been found to migrate and accumulate at the site of inflamed mucosa, resulting in microbial dysbiosis, intensified intestinal architectural damage, compromised resolution of inflammation, and an elevated risk of thrombosis[78]. On other hand, functional deficiency of neutrophils can accelerate disease progression, underscoring the multifaceted role of these granulocytes. In addition, our MR analysis found that the “neutrophil percentage of granulocyte” exposure displays a protective effect for allergic disease and rheumatoid arthritis. This phenotype is less straightforward to interpret unlike absolute counts and may be related to changed relative balance of granulocyte types between peripheral and disease injury sites.

Negative association of lymphocyte abundance with a number of IMD demonstrated in our MR analysis similarly requires a more fine-grained inspection, given disparate roles of various lymphocyte types (and their subtypes): T helper[79], T regulatory cells[80], memory T cells[81], B lymphocytes[82] etc. across the spectrum of inflammatory and autoimmune disease. Future larger GWAS studies of more fine-grained immunotypes will likely reveal new causal relationships for immune-mediated disease, such as shown recently for SLE[83]. Similarly, leveraging GWAS of specific disease subtypes as they become available will lead to more precise allocation of drug targets to conditions, which recognises their heterogeneity[84,85].

Having established putatively causal relationships between immune cell traits and IMD, we then proceeded onto gene-centric MR analysis prioritising drug targets using the intermediate trait approach. Following the paradigm of triangulation[86], we also corroborated and compared our findings with MR analyses utilising protein QTLs as exposures. The two approaches applied instruments derived from the same tissue – blood and focussed on targets with established roles in immunity, albeit largely non-overlapping between the two analysis sets, with only one-third shared. Altogether, we were able to track MR evidence for 548 drug targets at various stages in the development pipeline. Furthermore, we demonstrated the importance of utilizing multiple protein quantification studies, as unique signals were obtained from each. We also evaluated concordance across aptamer and antibody-based platforms to help derive consistent signals. Within intermediate and pQTL MR, we determined a substantial sharing of drug targets with MR evidence, especially for related atopic disease such as asthma, sinusitis and eczema, or different forms of inflammatory bowel disease: ulcerative colitis and Crohn’s disease. When compared, the two approaches yielded distinct disease sets with the highest number of drug targets with MR evidence, with only IBD overlapping across the top 4 in each method, thus providing complementary MR evidence.

Thanks to the utilization of the MR framework, our results are less likely to be affected by environmental confounding and reverse causation bias, factors that can impede causal inference in conventional epidemiological study designs. However, soundness of conclusions from our MR studies is related to fulfilment of main MR assumptions. We satisfied the *relevance* assumption by using a strict p-value threshold for exposure variants (p-value < 5 x 10^-8^) which resulted in strong instrumental variables (min F-stat=30). We used the colocalisation sensitivity method to test against the second assumption of *exchangeability.* Colocalisation compares the shape of pQTL or gene-specific intermediate trait association signal with outcome GWAS, which should reduce the number of linkage disequilibrium-related artefacts. Doubts remain around genes with coloc evidence located in the MHC region: *AGER*, *BRD2* and *LTA* which were found to associate with a number of IMD. The complex LD structure in the region, which extends beyond our colocalisation window[87] may have contributed to some false positives.

The colocalisation method employed by us requires the presence of just a single causal variant at the locus, unlike MR. This limitation was off-set by short computation time and lack of requirement for LD matrices for exposure and outcome GWAS[88]. This entails that small PP_H4_ values may not always represent evidence against colocalization, especially in cases where both PP_H3_ and PP_H4_ (both traits are associated, but with a different or shared causal variant, respectively) are small, indicating low power of analysis[67]. An additional measure undertaken against violation of the *exchangeability* assumption was to restrict our analysis to individuals of European descent, minimizing (but not eliminating[89]) the bias associated with population stratification. On the other hand, this may limit the generalizability of our findings to diverse human populations.

The *exclusion restriction* assumption was tested using the Egger intercept test, and it did not reveal much evidence for presence of horizontal pleiotropy, except for *IL5RA* pQTL. Nevertheless, this assumption cannot be definitively tested here due to the use of mostly single SNP instruments in intermediate trait and pQTL MR. However, restricting variants to *cis*-SNPs in proximity to the target gene is expected to limit bias from alternative pathways[15].

We encountered some limitations to our chosen study design. First of all, we were only able to find instruments for 548 out of 834 IMD drug targets (66%: 371 in intermediate MR and 361 in protein QTL) which have been deployed for preclinical and clinical investigations of IMD. Second of all, our MR analysis cannot instrument a drug that concurrently affects multiple parallel biological pathways. Multiple targets may be represented individually or in combination through a factorial MR approach[90] or meta-analysis[91], if only summary GWAS data are available. Since our analysis’ focus was on drug targets rather than mechanism of action for individual drugs, we took the former approach. In general, genetics-based methods such as MR cannot be informative about effectiveness of particular drug molecule chemistry[12]. Thirdly, our MR studies leverages only GWAS of disease incidence as there is a scarcity of genetic studies of disease progression[92]. Focusing solely on incidence may fail to capture genetic factors specifically associated with the severity, rapidity of onset and complications of the disease course.

Harnessing drugs which were already approved for use, we were able to quantify the sensitivity of both intermediate MR and pQTL approaches. Despite instrumenting a distinct set of targets, both methods arrived at similarly low average sensitivity (< 0.5). There is a host of possible reasons responsible for this result. In the intermediate trait MR, we may be lacking the most relevant immune cell phenotype to construct an instrumental variable with, such as counts of individual lymphocyte types. While blood is broadly of high relevance to all immune-mediated disease[4], for individual drug targets it may not always be the most biologically meaningful compartment to proxy drug’s mechanism of action[12]. That said, a number of organs and tissues release their proteins into the blood in health and disease, for example the complement proteins synthesised by the liver[93]. Plasma protein abundance and activation are influenced by post-translational mechanisms such as cleavage and secretion; these processes cannot be modelled using instruments derived simply from protein levels in the plasma. Context-specific QTLs from inflammatory states[94] or single-cell specific QTLs obscured by bulk tissue analysis[95,96] could act as more appropriate instruments in future studies.

Equally, pQTL analysis can be prone to false positives resulting from protein-altering variants which affect protein epitope; pQTLs detected in this case are going to reflect a mixture of changes due to the epitope effect and actual abundance[16]. Where proteins exist as inactive membrane-bound or circulating precursors as well as activated soluble forms, proteomic assays will be typically unable to clearly separate between different states. Such ambiguities makes interpreting the directionality of associations from MR problematic[94]. For instance, misaligned effect directionality relative to drug’s mechanism of action found in the pQTL MR for the association between TNFRSF1A and multiple sclerosis may stem from the exposure SNPs containing an epitope altering mutation. Furthermore, our pQTL MR results utilising non-PAV linked variants for two cytokine receptors: *interleukin 4 receptor* (*IL4R*) and *interleukin-2 receptor subunit alpha* (*IL2RA*) show opposite directions of effect to those established in the appropriate drug trials targeting asthma[97] and eczema[98,99], respectively.

We did also find strong evidence supporting truly directionally discordant effect of proteins whose pQTLs were not genetically linked to protein-altering variants. The role of individual proteins in IMD risk can be complex, with the same protein increasing risk for one condition while protecting against another. We replicated previous findings from Ferreira *et al.* (2013)[100], Rosa *et al.* (2019)[101] and Zhao *et al.* (2023)[94] for IL-6 receptor using cis-pQTL from across 4 pQTL cohorts, with opposing effects found on the risk of atopic disease and rheumatoid arthritis. We observed a similar phenomenon for BRD2 and CD40, with genetic predisposition to higher BRD2 increasing risk of multiple sclerosis but protecting against rheumatoid arthritis, and vice versa for CD40. This echoes real-world evidence from TNF-targeting therapies, which are efficacious for rheumatoid arthritis but not multiple sclerosis, where they may actually hasten disease progression[102]. As expected, we also uncovered disease-discordant significant effects using intermediate MR. *IL3*, *CSF2* and *FADS1* were found to display a positive association with asthma but negative with IBD, in the case of *CSF2* this could be also verified using pQTL instruments, with direction of effect in agreement. Such findings can diminish genetic support for drug repurposing across IMD.

## Conclusions

The present study utilized Mendelian Randomization (MR) to first clarify bidirectional genetic associations between peripheral immune cell phenotypes and a spectrum of IMD. We found a complex pattern of causal relationships, with elevated eosinophils associated with increased odds of several diseases and increased neutrophil percentage of granulocytes showing a protective effect for atopic conditions.

Intermediate trait Mendelian Randomization leveraging immune cell abundance instruments revealed 261 genes associated with immune-mediated diseases, with the highest count of hits for asthma, inflammatory bowel disease, eczema and chronic sinusitis. Colocalisation analysis provided further confirmation for 169 gene-disease pairs, with over 60% representing novel associations beyond existing drug indications. We then explored drug target prioritization utilising genetic associations of circulating protein concentrations as exposure. We compared results from different protein measurement technologies (SomaScan and Olink) which prioritized 284 proteins linked to diseases like inflammatory bowel disease and rheumatoid arthritis. Colocalisation analysis supported 83 protein-disease pairs, two-thirds of which were novel and may warrant experimental investigation.

Integrative analyses combining the intermediate trait and pQTL approaches showed relatively little overlap (13-39%), indicating their complementarity. Among shared signals, both validation for indications undergoing clinical trials and repurposing opportunities were revealed for: bempikibart and lusvertikimab (eczema – validation, asthma - repurposing) as well as lenzilumab, namilumab, plonmarlimab (asthma – validation, eczema – repurposing). New atopic indications for anakinra[103] could progress rapidly given that the drug is already approved.

Overall, multi-modal human genetics resources enable comprehensive evaluation of therapeutic candidates for immune-mediated disease. We highlight the importance of considering multiple data sources and methodologies in drug target prioritization for a more robust assessment. Given that ∼57% of pipeline compounds fail due to inadequate efficacy[104], genetically-informed indication selection holds certain promise to boost success rates.

## Data Availability

We accessed the following immune cell GWAS summary statistics via OpenGWAS (https://gwas.mrcieu.ac.uk/)[105]: ebi-a-GCST90002292, ebi-a-GCST004634, ebi-a-GCST90002380, ebi-a-GCST90002298, ebi-a-GCST004617, ebi-a-GCST90002382, ebi-a-GCST004614, ebi-a-GCST004608, ebi-a-GCST90002316, ebi-a-GCST90002389, ebi-a-GCST90002340, ebi-a-GCST90002394, ebi-a-GCST004626, ebi-a-GCST90002351, ebi-a-GCST004623, ebi-a-GCST90002399, ebi-a-GCST004620, ebi-a-GCST004624, ebi-a-GCST004613, ebi-a-GCST90002374. We accessed the following immune-mediated disease GWAS summary statistics via OpenGWAS (https://gwas.mrcieu.ac.uk/)[105]: ebi-a-GCST90014325, ebi-a-GCST006862, ebi-a-GCST004132, ebi-a-GCST004131, ieu-b-18, ebi-a-GCST90019016, ebi-a-GCST002318, ebi-a-GCST003156, ebi-a-GCST010681, ebi-a-GCST004133. We obtained the following immune-mediated disease GWAS summary statistics from FinnGen ver. 9 (https://www.finngen.fi/en/access_results)[39]: M13_ANKYLOSPON, J10_CHRONSINUSITIS, L12_ATOPIC and NHGRI-EBI GWAS Catalog (http://www.ebi.ac.uk/gwas)[106]: GCST90244787, GCST90027899, GCST90010715. UKBB pQTL dataset[55] was accessed via https://metabolomips.org/ukbbpgwas/, deCODE pQTL dataset[53] was accessed via https://www.decode.com/summarydata/, ARIC pQTL dataset[52] was accessed via http://nilanjanchatterjeelab.org/pwas/.

## Ethics approval and consent to participate

The article uses previously published GWAS summary statistics. No separate ethical approval is required for this study. All subjects provided informed consent in the original studies.

## Author contributions

MKS: Conceptualization, Formal analysis, Visualization, Writing – original draft, Writing – review and editing

TG: Supervision, Writing – review and editing, Funding acquisition, Resources

