## Supplemental Figures for "Integrative Mendelian Randomization approaches for therapeutic target prioritisation in immune-mediated diseases"

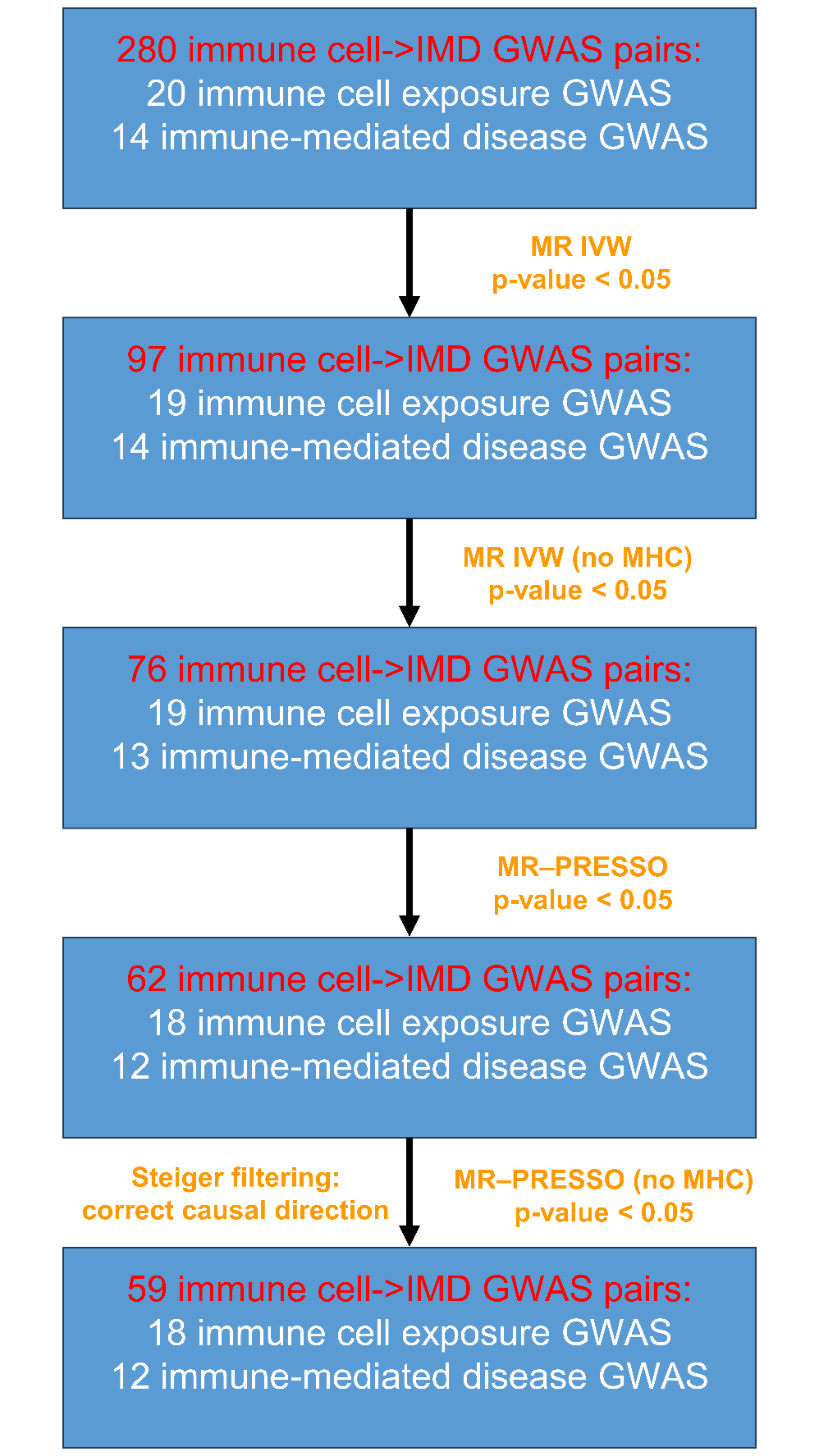

**Supplementary Figure 1.**  Filtering of MR results for the effect of immune cells on immune-mediated disease (IMD) to arrive at the final set of 59 exposure-outcome associations deemed high-confidence. IVW – inverse-variance weighted; no MHC – MR analyses excluding the MHC genomic region.

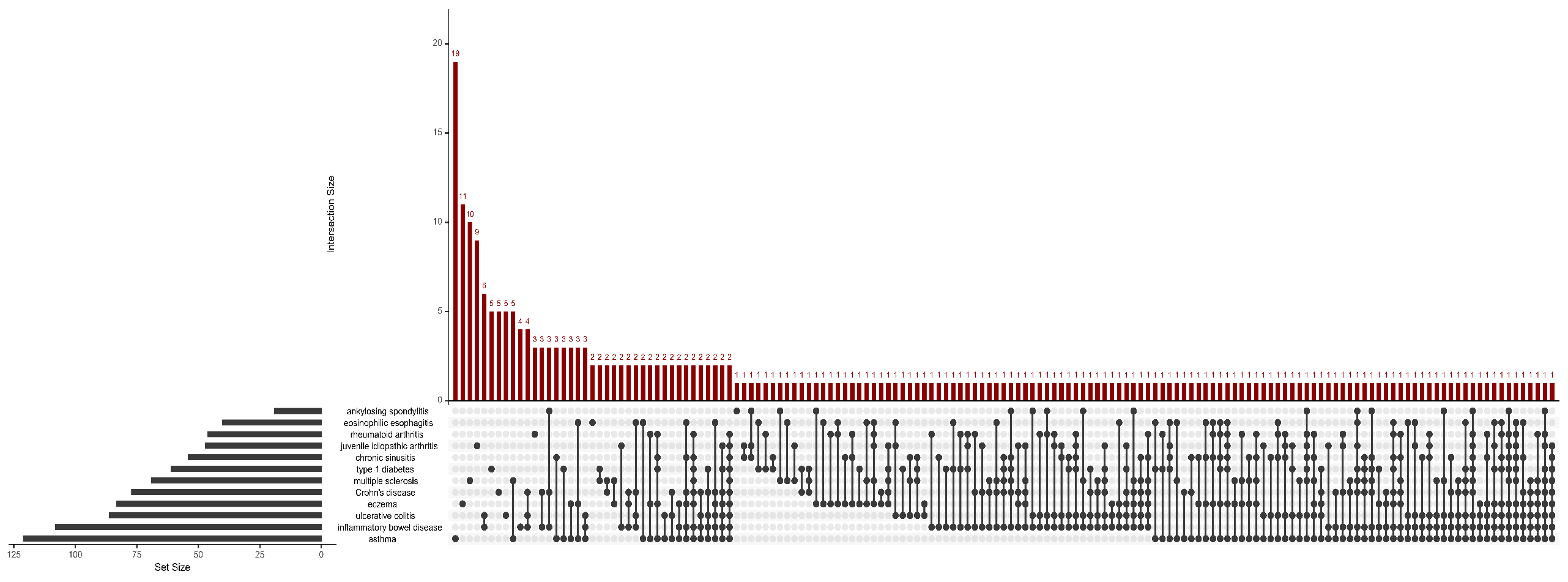

**Supplementary Figure 2.** UpSet plot showing overlap of drug targets across IMD in the intermediate trait MR analyses. We included only nominally significant (p-value < 0.05) MR associations.

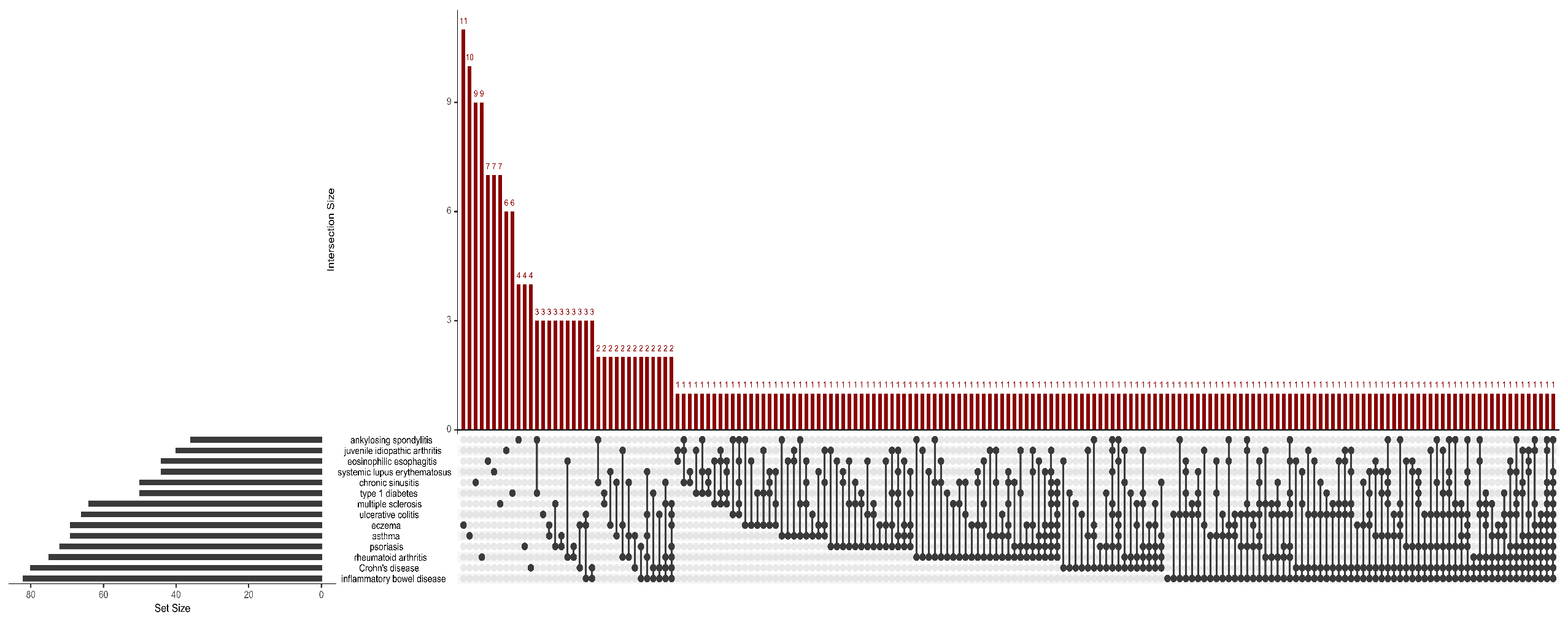

**Supplementary Figure 3**. UpSet plot showing overlap of drug targets across IMD in the pQTL MR analyses. We included only nominally significant (p-value < 0.05) MR associations.

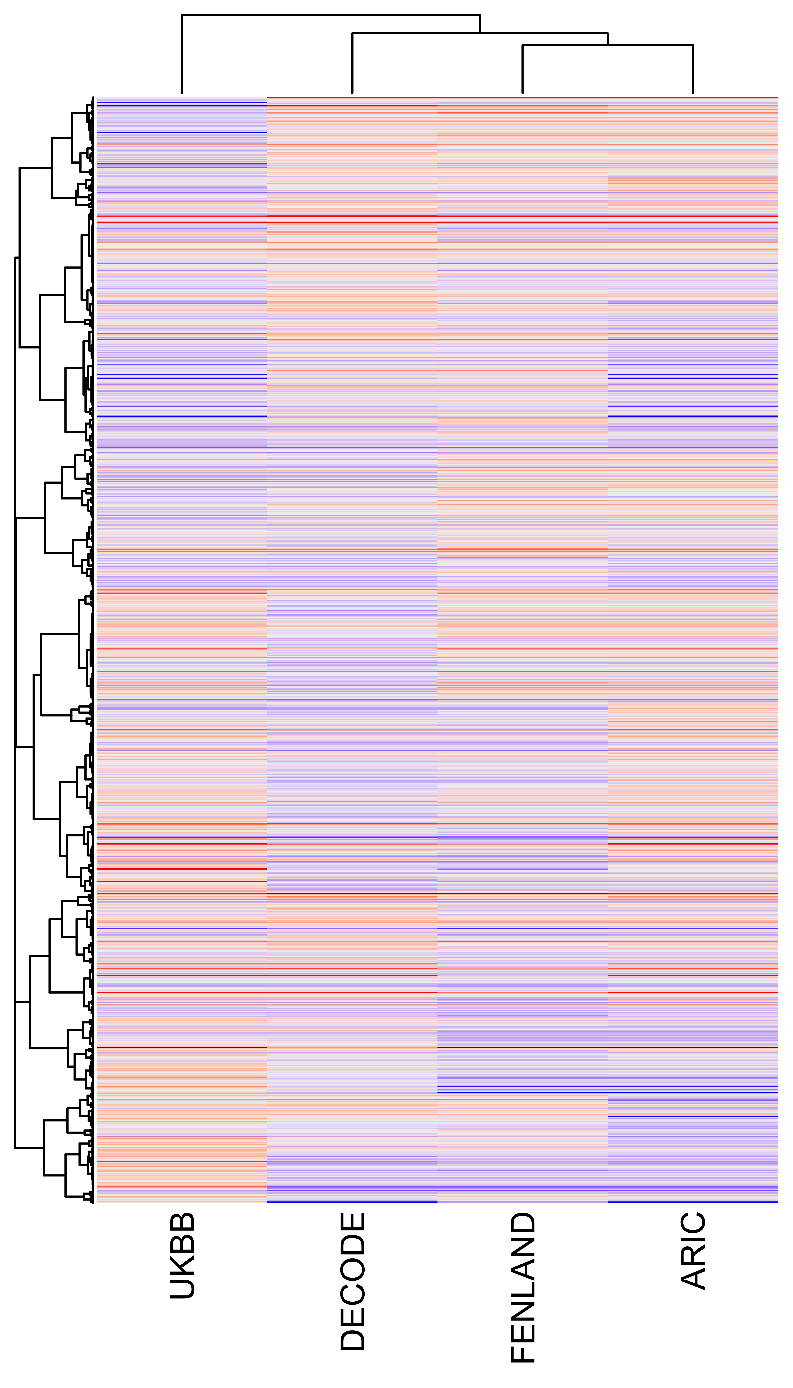

**Supplementary Figure 4**. Hierarchically clustered z-scores of MR estimates across pQTL sources. Distance metric used was Pearson's *r* and clustering algorithm was complete-linkage.

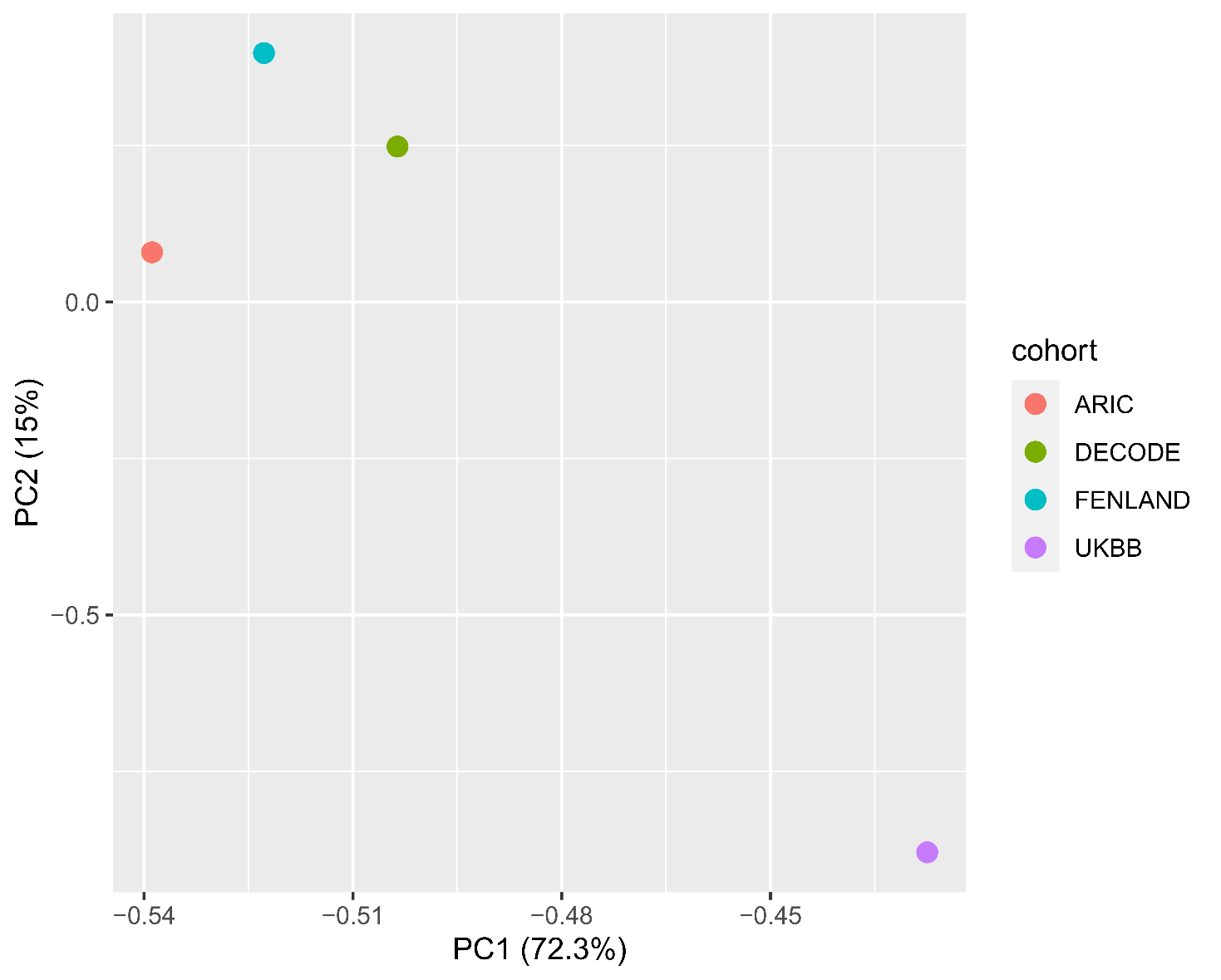

**Supplementary Figure 5**. Principal component analysis (PCA) of z-scores of MR estimates across pQTL sources. PC1 and PC2 correspond to 72.3% and 15% of explained variance, respectively.

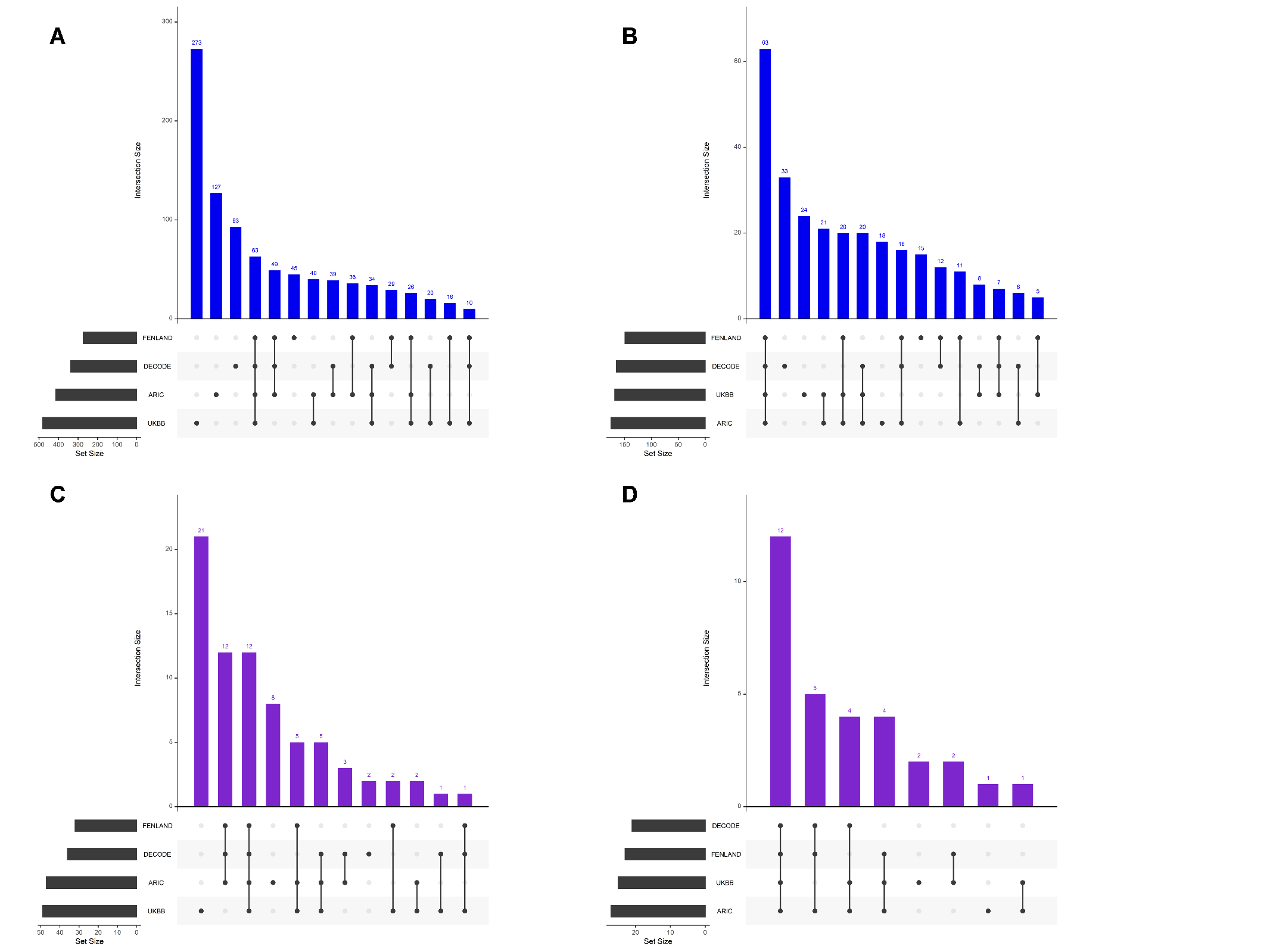

**Supplementary Figure 6.** UpSet plots showing protein-IMD association overlap across pQTL sources. **A**) nominal p-value threshold met, including all tested proteins with a suitable instrument; **B**) nominal p-value threshold met, including only overlapping target proteins among pQTL instrument sources; **C**) Bonferroni-corrected p-value threshold (p-value < 10^-7^)* met in either set, including all tested proteins with a suitable instrument; **D**) Bonferroni-corrected p-value threshold met in either set, including only overlapping target proteins among pQTL instrument sources.

*Bonferroni-corrected threshold equalled: 0.05 / (number of proteins tested x number of outcomes tested)
